# A benign control-calibrated multiparametric functional assay supports *CACNA1A* variant assessment

**DOI:** 10.64898/2026.10.12.26365280

**Authors:** Chek-Ying Tan, Neven Maksemous, Joanne G. Ma, Sangavi Sivagnanasundram, Qianyi Shen, Shrey Sharma, Himanshu Goel, Manny Jacobs, Zornitza Stark, Kavitha Kothur, Lyn Griffiths, Jamie I. Vandenberg, Chai-Ann Ng

## Abstract

*CACNA1A*-related disorders arise from altered function of the P/Q-type calcium channel Cav2.1 and encompass a broad spectrum of neurological phenotypes, including developmental and epileptic encephalopathy, familial hemiplegic migraine type 1 and episodic ataxia type 2. Although loss-of-function and gain-of-function mechanisms have both been implicated, interpretation of rare *CACNA1A* missense variants remains challenging because clinical presentation is heterogeneous and many variants identified through diagnostic sequencing remain classified as variants of uncertain significance. To generate clinically interpretable functional evidence, we developed a calibrated high-throughput automated patch-clamp assay to assess the *CACNA1A* variants.

Twenty-six curated benign/likely benign and 30 curated pathogenic/likely pathogenic missense variants were expressed in Flp-In HEK293 cells with *CACNB4* and *CACNA2D1*. We quantified current density, Δ*V*_0.5_ activation, Δ*V*_0.5_ inactivation and recovery from inactivation (*T*_0.5_ RFI), and used benign controls to define parameter-specific normal ranges with ±3 Z-score thresholds. This multi-parameter framework classified all benign controls as functionally normal and 29 of 30 pathogenic controls as abnormal, yielding 100% specificity and 96.7% sensitivity. Current density identified 19 pathogenic controls with loss of function, while gating and recovery from inactivation identified additional abnormal variants with preserved or partially preserved current density. The assay generated likelihood ratios of 0.033 for normal function and 25.13 for abnormal function, supporting BS3 and PS3 functional evidence, respectively.

Application to 23 *CACNA1A* variants identified in Australian patients showed abnormal function in all four variants already classified as pathogenic or likely pathogenic. Among 19 variants of uncertain significance, 11 were functionally abnormal and ten were reclassified as likely pathogenic after integration of PS3 evidence with other ACMG/AMP criteria. These data show that *CACNA1A* missense variants can produce loss-of-function, gain-of-function or mixed effects, and that multi-parameter functional assessment is required for accurate interpretation. This calibrated automated patch-clamp framework provides clinically interpretable evidence for refining *CACNA1A* variant classification in diagnostic practice.

## Introduction

The voltage-gated calcium channel Cav2.1, which mediates P/Q type Ca^2+^ current, is essential for neurotransmitter release and thereby regulates neuronal excitability and synaptic transmission.^1–4^ Cav2.1 is widely expressed in presynaptic terminals throughout the brain, including the thalamus, cortex and hippocampus, and is particularly abundant in cerebellar Purkinje cells and granule cells, where it has a central role in motor coordination and balance.^5–7^ Pathogenic variants in *CACNA1A*, which encodes the pore-forming α1 subunit of Cav2.1, cause a broad spectrum of neurological, neuropsychiatric, and neuromuscular disorders.^8^ Missense *CACNA1A* variants are classically associated with developmental and epileptic encephalopathy (DEE), familial hemiplegic migraine type 1 (FHM1) and episodic ataxia type 2 (EA2), which often have distinct but overlapping ages of onset and clinical trajectories. DEE, typically presenting in infancy with seizures, developmental delay, intellectual disability and ataxia, can be caused by both loss-of-function (LOF) and gain-of-function (GOF) *CACNA1A* variants.^9,10^ FHM1, a rare form of recurrent migraine characterised by aura and transient motor weakness, is typically caused by GOF *CACNA1A* variants.^11,12^ EA2, one of the most common forms of episodic ataxia subtype,^13^ characterised by recurrent attacks of ataxia, nystagmus and vertigo, often triggered by stress or fatigue in adolescence, is predominantly associated with LOF *CACNA1A* variants.^14,15^ Despite these broad genotype-phenotype associations, *CACNA1A* is highly pleiotropic, and individual variants may produce mixed or variable phenotypes, which makes diagnosis and management difficult.^6,16^ An accurate molecular diagnosis is also important for patient management as different classes of *CACNA1A* variants will likely require different therapeutic approaches. For example, FHM1 patients usually respond well to verapamil, flunarizine and topiramate, consistent with variants that cause a GOF.^11,17–19^ In contrast, patients with EA2, who typically have *CACNA1A* LOF variants, benefit from acetazolamide or 4-aminopyridine.^14,20^ However, treatment response is variable, and this may partly reflect differences in the underlying functional defect caused by the individual variant.

*CACNA1A* is a large gene in which rare missense variants are common in the general population, making it difficult to distinguish benign from pathogenic changes. The increasing use of next-generation sequencing, driven in part by falling sequencing costs, has further accelerated the discovery of rare variants. Consequently, more than 2000 missense *CACNA1A* variants have been reported in ClinVar, of which 74% are classified as variants of uncertain significance (VUS). Such unresolved findings can result in diagnostic uncertainty, limit access to targeted treatments or family screening, and impose a substantial psychological burden on patients, families and clinicians. Thus, there is a critical need for robust strategies to support variant reclassification. Under the American College of Medical Genetics and Genomics and the Association for Molecular Pathology (ACMG/AMP) variant classification guidelines, multiple lines of evidence are required to reach a pathogenic or likely pathogenic classification.^21^ Functional evidence from well-established assays can provide strong evidence for variant classification, but most functional studies use only wild type (WT) as a sole or a small number of population variants as reference, rather than the standardised framework of systematically curated benign and pathogenic controls recommended by ClinGen;^22^ these assays therefore have limited utility for variant classification in a diagnostic setting.

Pathogenic *CACNA1A* variants can disrupt Cav2.1 function through reducing membrane abundance and/or altering channel gating.^23,24^ Many clinically observed missense variants result in both trafficking and gating defects, producing mixed phenotypes. Here, we developed a calibrated multi-parameter functional assay using a systematically curated panel of 26 known benign and 30 known pathogenic controls, to define an objective threshold for abnormal function, enabling reclassification of nine abnormal *CACNA1A* VUS in Australian patients.

## Method

The experimental design and workflow of the calibrated *CACNA1A* automated patch clamp assay are illustrated in Fig. 1.

**Figure 1.**
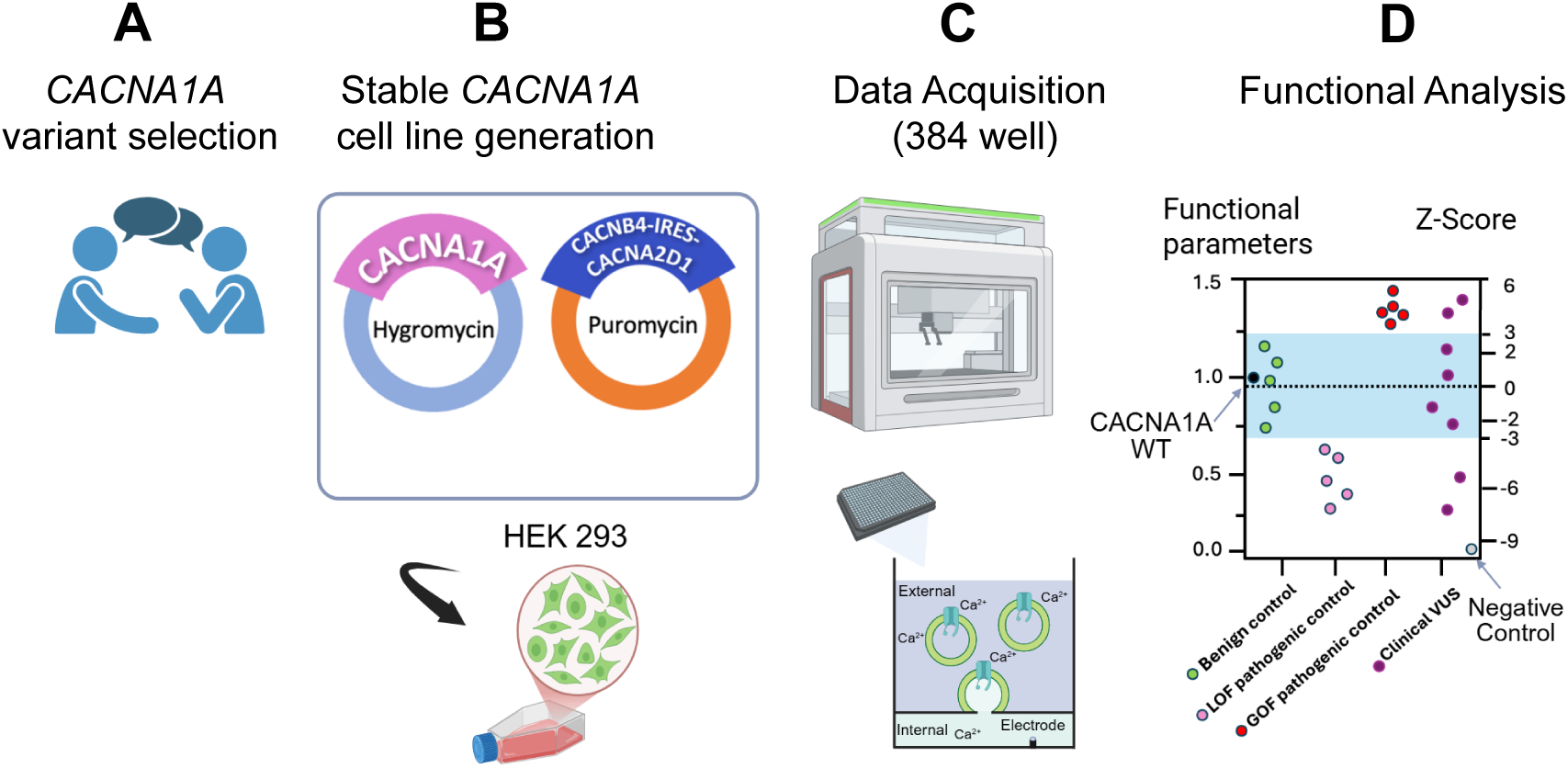
Overview of the calibrated *CACNA1A* automated patch-clamp assay. (A) Benign and pathogenic *CACNA1A* calibration variants were curated according to ACMG/AMP criteria, and variant-containing plasmids were generated by GenScript Inc. (B) Stable Flp-In HEK293 cell lines were generated using a double-Flp-In strategy to express the pore-forming Cav2.1 α1 subunit encoded by *CACNA1A*, together with the auxiliary subunits *CACNB4* and *CACNA2D1*.^26^ Cells successfully incorporating the constructs were selected using blasticidin, hygromycin and puromycin. (C) Ca^2+^ currents from WT, benign, pathogenic and clinical *CACNA1A* variant cell lines were recorded using the Nanion SyncroPatch 384 automated patch-clamp platform. (D) Functional readouts including current density, voltage dependence of activation, voltage dependence of inactivation and recovery from inactivation were converted to benign control-calibrated Z-scores to distinguish functionally normal from abnormal *CACNA1A* variants.

### CACNA1A, CACNB4, and CACNA2D1 transcripts

Variants were annotated and experimentally studied using the MANE Select *CACNA1A* transcript ENST00000360228.11 (NM_001127222.2), co-expressed with *CACNB4* ENST00000539935.7 (NM_000726.5) and *CACNA2D1* ENST00000356860.8 (NM_000722.4), the most conserved and highly expressed transcripts in the cerebellum.

### Selection and curation of *CACNA1A* control variants

The *CACNA1A* control variants were curated and classified according to the American College of Medical Genetics and Genomics and Association for Molecular Pathology (ACMG/AMP) guidelines.^21^ All variants were analysed in the context of the GRCh38 (hg38) genome build. Population frequency data were obtained from gnomAD v4.1.1 (genomes dataset), excluding the Amish cohort (allele count <1,000). A minor allele frequency (MAF) threshold of 1 in 10,000 was applied, consistent with the estimated prevalence of dominantly inherited hemiplegic migraine;^25^ variants exceeding this threshold were assigned the BS1 criterion. Criterion BS2 was applied at either strong or supporting strength based on the occurrence of homozygous or heterozygous variants in healthy individuals. Pathogenic evidence included PS2 or PM6 for confirmed or assumed *de novo* variants, applied according to ClinGen Sequence Variant Interpretation (SVI) Working Group recommendations (v1.1). PS4 (moderate strength) was assigned to variants reported in multiple unrelated affected individuals and absent from population controls. PM2 (supporting) was applied to variants that were absent or rare in gnomAD. PP3 was assigned using ClinGen-calibrated AlphaMissense predictions,^26^ with evidence strength capped at moderate. Functional evidence was not applied to any control variant during classification. The final curated dataset comprised 26 benign/likely benign (B/LB) and 30 pathogenic/likely pathogenic (P/LP) variant controls (Supplementary Table 1).

### Cloning of *CACNA1A* constructs and generation of *CACNA1A* variant cell lines

Wild type *CACNA1A* cDNA in pcDNA^TM^/FRT/TO was designed to have seven segments separated by restriction sites (NheI, NsiI, StuI, AscI, KpnI, ClaI, BstEII, BamHI) to facilitate subcloning of variants by GenScript Inc (Piscataway, USA). The stable transfection of three genes encoding the Cav2.1 complex in Flp-in HEK293 cells was achieved using to the double-Flp-in method described by Jensen *et al*.^27^ The generation of the *CACNA1A* variants, restriction digest, subcloning into the pcDNA^TM^5/FRT/TO vector, and confirmatory Sanger sequencing were completed by GenScript Inc (Piscataway, USA). The first plasmid contains a copy of the *CACNA1A* variant, while the second plasmid contains *CACNB4* and *CACNA2D1*, separated by an internal ribosomal entry site (IRES) element to provide the essential subunits for optimal Cav2.1 function. Plasmids (100 ng of each plasmid) were transfected with 1800 ng pOG44 Flp-Recombinase Expression Vector (Thermo Fisher Scientific, cat. # V600520) into Flp-in HEK 293 (Thermo Fisher Scientific, cat #R78007) parental cells using Lipofectamine 3000 (Thermo Fisher Scientific, cat. # L3000015) following manufacturer’s instructions. Stable Flp-in cell lines were selected using BHP-DMEM media (Thermo Fisher Scientific, cat. # 10566024) supplemented with 10% fetal bovine serum and an antibiotic cocktail containing 10 μg/mL Blasticidin (InvivoGen, cat. #ant-bl-1), 100 μg/mL Hygromycin (Thermo Fisher Scientific, cat. # 10687010), and 0.25 μg/mL Puromycin (Gibco, cat. # A11138-03). Cells were kept at 37 °C in a humidified incubator with 5% CO_2_. For electrophysiology experiments, the Cav2.1 protein complex expression was induced with 200 ng/mL doxycycline (Sigma-Aldrich, cat. # D9891) for 48 hours prior to experimentation.

### *CACNA1A* automated patch clamp assay

Ca^2+^ currents were recorded at room temperature using the SyncroPatch 384 (Nanion Technologies, Munich, Germany), an automated patch clamp instrument with 384 recording channels. *CACNA1A* WT, benign (B) and pathogenic (P) control variants, and negative control cell lines were loaded into the Nanion-S-type chips (Nanion Technologies, Munich, Germany). The internal solution contained (in mmol/L) 10 NaCl, 10 CsCl, 110 CsF, 10 HEPES, 10 EGTA (pH 7.2 with CsOH). The external solution contained (in mmol/L) 20 NaCl, 5 KCl, 1 MgCl_2_, 2 CaCl_2_, 120 TEACl, 10 HEPES, 5 Glucose (pH 7.4 with NaOH). Corrections for liquid junction potential was −12.8 mV. Whole-cell currents were recorded at 2-10 kHz sampling frequency.

### Recording protocols and data analysis

Cell peak current density (pA/pF), calculated as peak current (pA), measured recorded using a 25 ms depolarising test pulse to zero mV, from a holding period of −80 mV, divided by cell capacitance (pF).

Steady-state activation was recorded using 25 ms depolarising step pulses to voltages ranging from −50 mV to +40mV in 10 mV increments, from a holding period of −120 mV to ensure all channels are in their resting state before the depolarizing pulses are applied. Current was converted into conductance according to the equation:

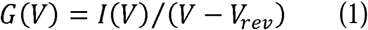

where *G*(*V*) is conductance at the test potential *V*,*I*(*V*) is amplitude of the current measured at test potential *V,V* is the membrane voltage, and _*Vrev*_ is the reversal potential.

Steady-state inactivation was measured from peak currents recorded using a 20 ms depolarising pulse to zero mV, preceded by 750 ms-long conditioning pulses (ranging from −90 mV to +40 mV in 10 mV increments). The inter-pulse holding potential was −120 mV to ensure uniform resting states.

Steady-state activation and inactivation data were fitted using a Boltzmann equation:

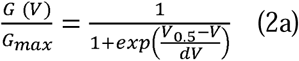

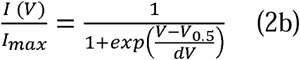

where *V* is membrane voltage, *V*_0.5_ is membrane voltage at which half-maximal activation (*G*) or half-maximal inactivation (*I*) is reached, *I*_max_ is maximal current amplitude where *V* is membrane voltage, *V*_0.5_ is membrane voltage at which half-maximal activation (*G*) or half-maximal inactivation (*I*) is reached, *I*_max_ is maximal current amplitude and is the slope factor.

Recovery from inactivation (RFI) was assessed using a double-pulse protocol. Both the pre-pulse and test pulse were depolarising pulses to zero mV for two seconds, separated by variable inter-pulse intervals (ranging from 50 ms to 15 seconds) at a holding potential of −120 mV. The time interval between sweeps was set to 28 seconds to ensure maximum recovery between sweeps. All traces were normalised to the maximum current during the pre-pulse for each sweep. The peak current from the test pulse was plotted as a percentage of maximum pre-pulse current vs. repolarisation time. The envelope of peak current was fit with a two-phase exponential equation to obtain the time for half recovery from inactivation (*T*_0.5_). The envelope of peak current was fit with a two-phase exponential equation to obtain the time for half recovery from inactivation (*T*_0.5_ RFI).

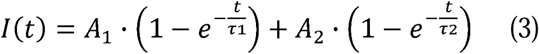

Where *I*(*t*) is the current amplitude at a recovery interval *t, A_1_* and *A_2_*are the amplitudes of the fast and slow components, τ_1_ and τ_2_ are the time constants for the fast and slow phases of recovery, respectively.

Each *CACNA1A* variant was assayed on four separate plates. Two technical replicates were performed during the same-day experiment, and this process was repeated using cells from a different frozen stock for the same construct. WT control lines were included on every run and peak current densities, *V*_0.5_ act, *V*_0.5_ inact, and *T*_0.5_ RFI measurements for *CACNA1A* variants were normalised to WT mean recorded on the same plate to minimise plate-to-plate variations. Recordings were analysed using the Data Control 384 software (Nanion Technologies, Munich, Germany).

### Quality control and sample size

The quality control for all data includes seal resistance > 500 MΩ, capacitance (3-50 pF), series resistance < 15 MΩ, and leak corrected < ±40 pA at −120 mV. Additional QC parameters were applied for gating measurements, including voltage errors due to uncompensated series resistance < 10 mV, Boltzmann fit had R^2^ > 0.99, reversal potential 45-60 mV, peak current amplitude −300 pA to −2000 pA, and series resistance < 10 MΩ. For RFI analysis, data were excluded if double exponential fit R^2^< 0.97 or if recovery percentage was > 120%.

Power calculations were performed using WT single-cell recordings that passed quality-control criteria. For each electrophysiological parameter, the minimum number of cells required was estimated to detect a shift equivalent to the benign-control three Z-score threshold at 90% power with α = 0.05.

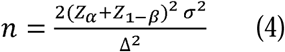

Where *n* is the required sample size required per group, _*Za*_ is the constant for a two-sided α error of 0.05, *z*_1–β_ is the constant corresponding to a statistical power of 90%, σ is the standard deviation, Δ is the absolute difference between the two means.

### Z-score and likelihood ratio calculations

For each functional property, normalised means of 26 benign controls were used to establish the parameter-specific normal functional range. Z-scores, representing the number of standard deviations from the benign-control mean, were calculated as follows:

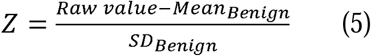

The ACMG/AMP evidence level was derived from the likelihood ratio as below:

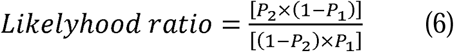

Where *P*_1_ is the prior probability of pathogenicity and *P*_2_ is the posterior probability after the assay result.

## Results

### Topographic distribution of benign and pathogenic Cav2.1 variants across channel domains

*CACNA1A* encodes the Cav2.1 α1 subunit, a large membrane protein composed of four homologous repeats (I–IV), each containing six transmembrane segments (S1–S6), with S4 segments forming the principal voltage-sensors and S5–S6 segments with pore loops forming the ion-conducting pore.^28^ A total of 26 benign and 30 pathogenic *CACNA1A* missense variants were curated without using functional data. The 26 benign controls were all located in the intracellular regions, including the cytosolic N- and C-terminal domains and intracellular linkers, whereas pathogenic controls were enriched in extracellular, transmembrane and pore-forming regions, including S4 voltage-sensing segments, S5–S6 pore regions and adjacent extracellular loops (Fig. 2A).

**Figure 2.**
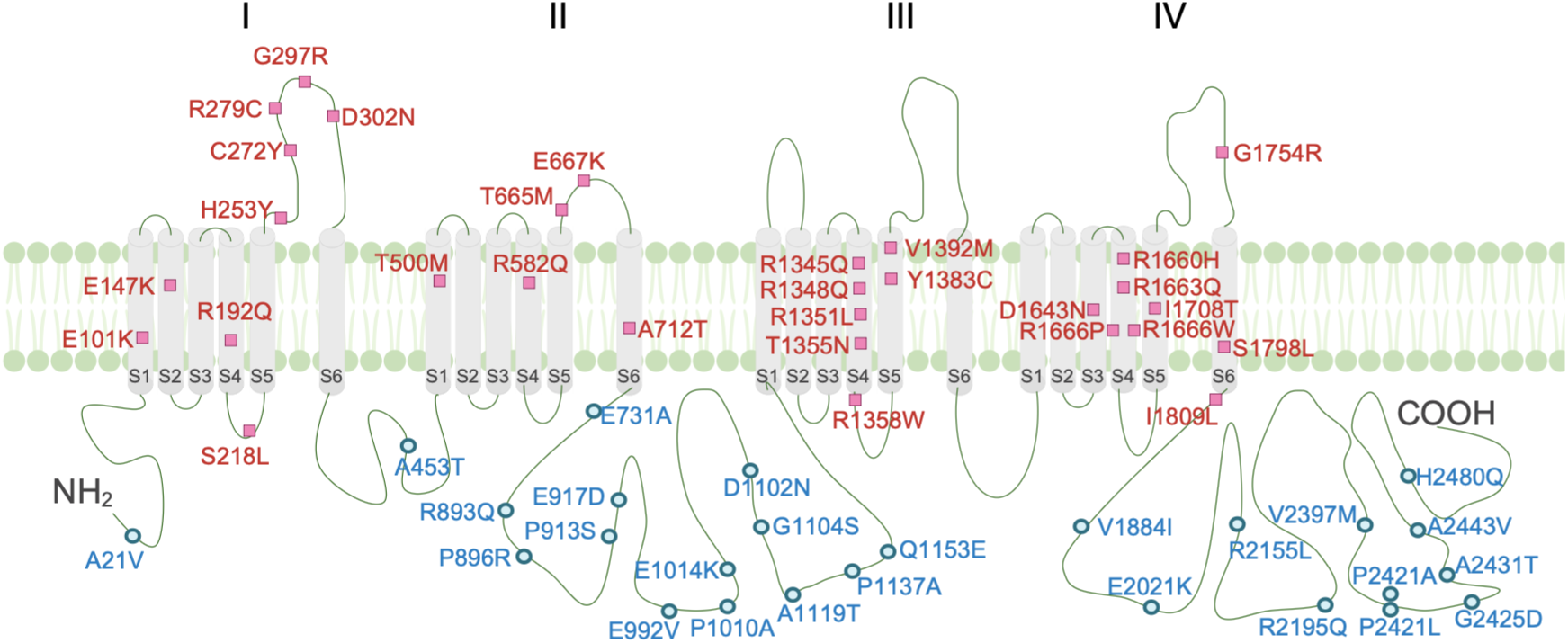
Membrane topology and locations of benign and pathogenic *CACNA1A* calibration variants. Domains I–IV correspond to the four homologous transmembrane domains of the Cav2.1 α1A subunit. Each domain contains six transmembrane segments (S1– S6), with the S4 segments serving as voltage sensors. Benign control variants are indicated by blue circles and pathogenic control variants by red squares. Variants are distributed throughout the channel, including the transmembrane domains, intracellular linkers, N-terminus and C-terminus. A total of 26 missense benign (blue) and 30 pathogenic control variants (red) were included.

### Current density as the primary discriminator for LOF

Current density, measured from the peak current during a 25 ms depolarisation step to zero mV from a holding potential of −80 mV, provides measurement of functional channels expression at the plasma membrane (Fig. 3A). The distribution of WT Cav2.1 current density from 1040 cells showed a positively skewed distribution (inset of Fig. 3A). To achieve a normal distribution, a Y^0.75^ transformation was applied followed by WT normalisation. From the distribution in Fig. 3A, a minimum of 28 cells was required to detect a three Z-score difference in current density at 90% power with a 95% confidence interval (CI).

**Figure 3.**
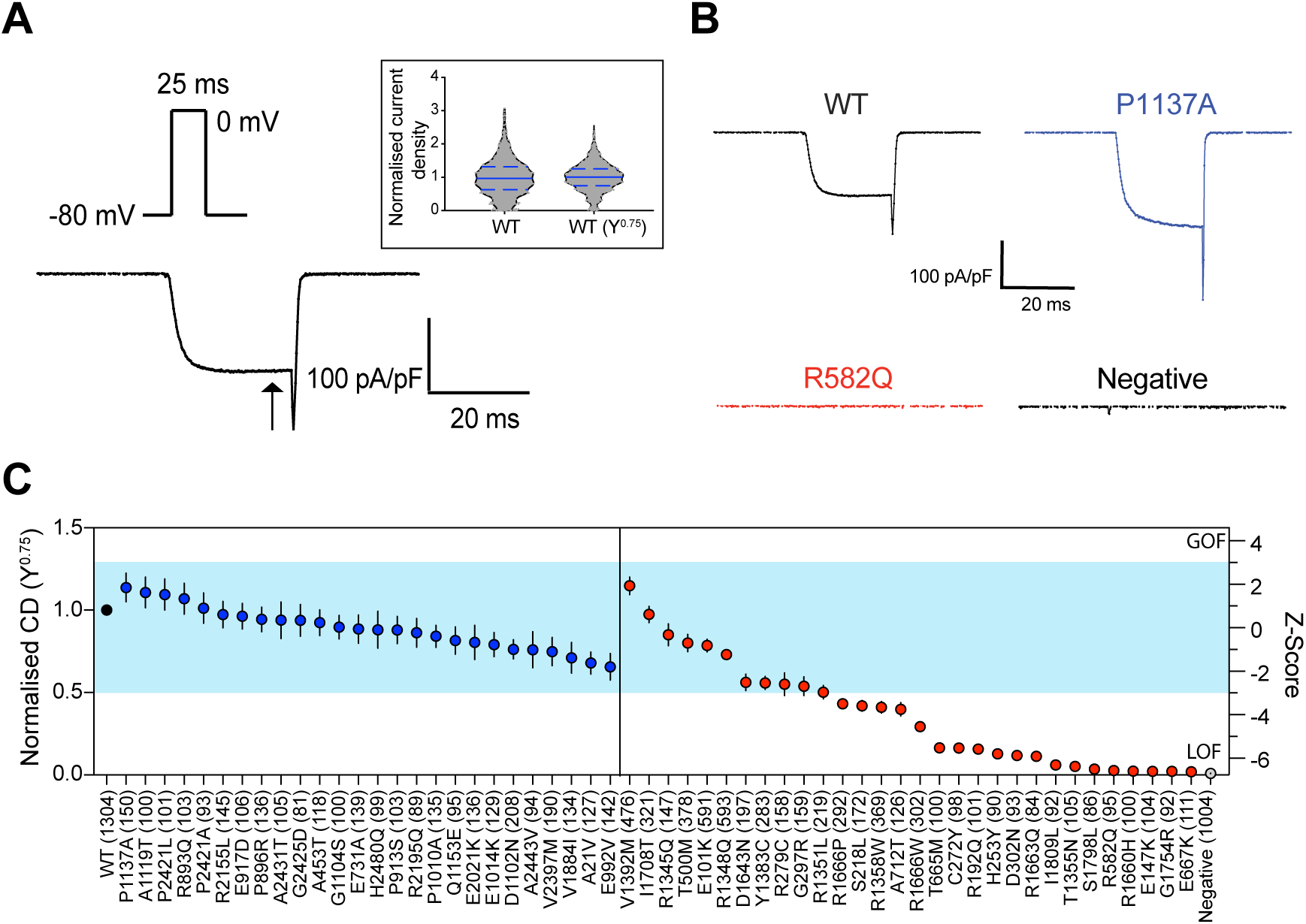
Benign control-calibrated current density analysis identifies LOF *CACNA1A* variants. (A) Representative whole-cell Ca^2+^ current trace recorded from a single cell expressing WT *CACNA1A* in response to a 25 ms depolarising step from −80 mV to 0 mV. The inset shows violin plots of current density values from 1040 cells recorded on WT-only plates, before and after Y^0.75^ transformation and normalisation to the WT mean. Median values and first and third quartiles are indicated by blue lines. (B) Representative current traces are shown for WT *CACNA1A* (−134 pA/pF, black), the benign control P1137A (−199.1 pA/pF, blue), the LOF pathogenic control R582Q (−1.09 pA/pF, red), and a non-expressing negative control cell (−0.73 pA/pF, grey). (C) Using the Z-score grading system, all 26 benign *CACNA1A* control variants fell within the ±3 Z-score normal range (blue region), whereas 19 of 30 pathogenic control variants had abnormal current density outside the blue normal range, consistent with LOF. Numbers of cells are shown in parentheses; WT n includes all WT cells recorded, comprising WT-only plates and the WT controls included on every variant plate. Data are presented as mean ± 95% CI.

Representative current traces for a known benign control (P1137A), a known pathogenic control (R582Q), and a negative control cell are shown in Fig. 3B. The mean and standard deviation of the normalised current density based on the distribution of 26 benign control variants was 0.89 ± 0.131 (SD), which is the blue shaded box in Fig. 3C. Nineteen of 30 pathogenic controls have Z-scores smaller than −3, indicating a LOF phenotype. The 11 pathogenic controls for which current density did not identify abnormal functions were E101K, R279C, G297R, T500M, R1345Q, R1348Q, R1351L, Y1383C, V1392M, D1643N and I1708T (Supplementary Table 2).

### Altered voltage-dependence of activation in pathogenic variants with normal current density

Cav2.1 variants can also alter function by affecting channel gating. There were 16 variants with sufficient current density (Z > −5) to enable analysis of gating (Supplementary Table 2). Representative WT current traces recorded during 25-ms depolarisation steps from −50 to +40 mV in 10 mV increments, from a holding potential of −120 mV, and the corresponding I−V relationship are shown in Fig. 4A. The corresponding conductance−voltage curves are fitted with a Boltzmann function, giving a WT midpoint of activation (*V*_0.5_ act) of –4.8 mV (Fig. 4B). For comparison, conductance−voltage curves are also shown for a benign variant R2155L (*V*_0.5_ act = –7 mV) and two pathogenic variants: E101K (*V*_0.5_ act = −20.8 mV), showing a hyperpolarising shift, and D1643N (*V*_0.5_ act = −0.43 mV), showing a depolarising shift (Fig. 4B).

**Figure 4.**
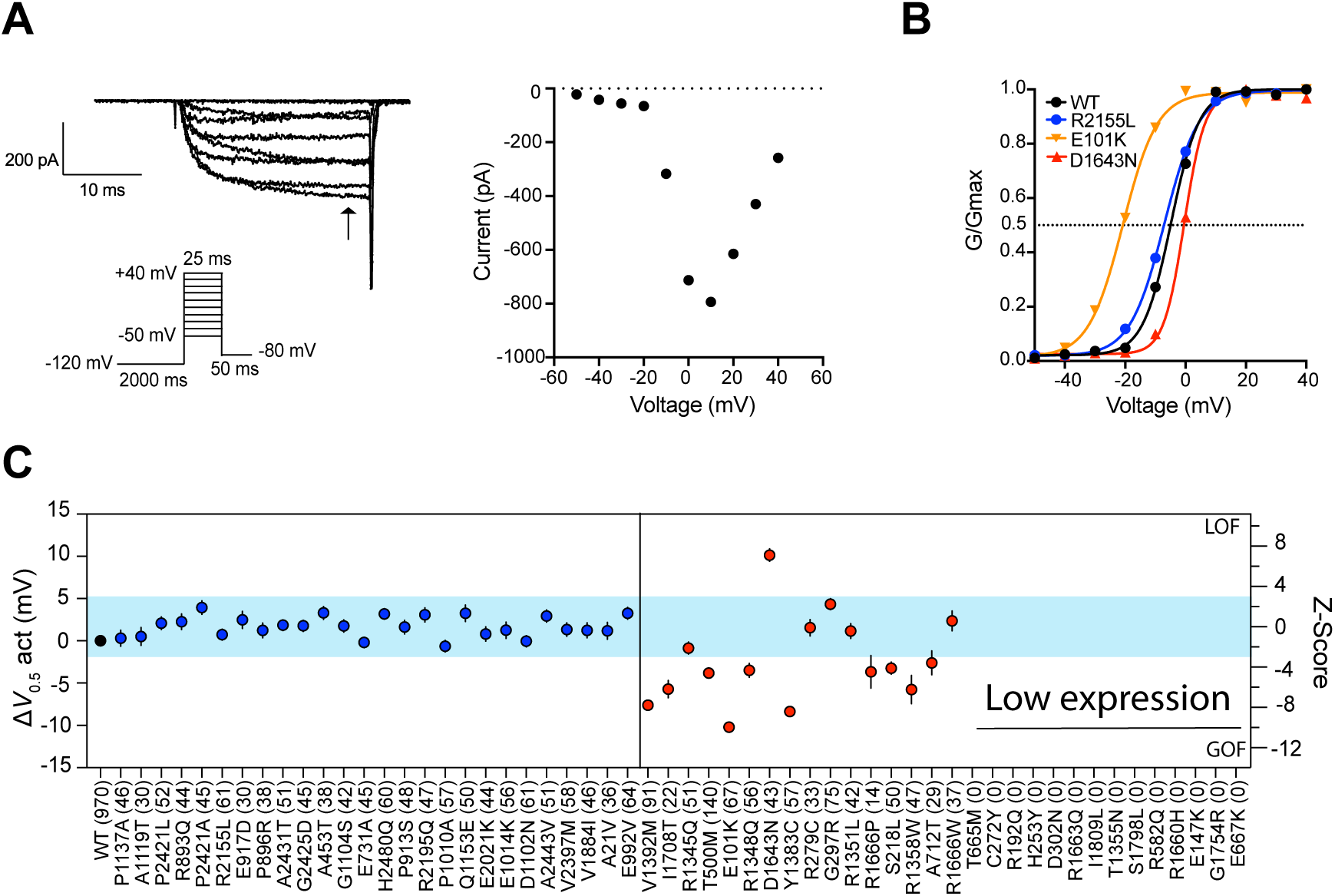
Benign control-calibrated assessment of voltage dependence of activation identifies GOF and LOF *CACNA1A* variants. (A) Representative whole-cell Ca^2+^ currents recorded from a WT HEK293 cell expressing *CACNA1A*. **Left:** family of current traces evoked by 25 ms depolarising steps from −50 mV to +40 mV in 10 mV increments from a holding potential of −120 mV. **Right:** plot of peak current at each test potential. (B) Current amplitudes from panel A were converted to conductance and normalised to maximal conductance. Solid lines represent Boltzmann fits to the experimental data. Example conductance-voltage curves are shown for WT *CACNA1A* (*V*_0.5_ act = −4.8 mV, black), the benign control R2155L (*V*_0.5_ act = −7 mV, blue), the pathogenic control D1643N (*V*_0.5_ act = −0.43 mV), which shows a depolarising shift consistent with LOF, and the pathogenic control E101K (*V*_0.5_ act = −20.8 mV), which shows a hyperpolarising shift consistent with GOF. (C) The *V*_0.5_ act of each variant was averaged and subtracted from the WT mean recorded on the same plate to calculate Δ*V*_0.5_ act. Numbers of cells analysed for each variant are shown in parentheses. From analysis of WT cells, a minimum of 11 cells was required to detect a three Z-score difference in *V*_0.5_ act at 90% power with a 95% CI. Using the Z-score grading system, Δ*V*_0.5_ act values for all 26 benign *CACNA1A* control variants fell within the ±3 Z-score normal range. Of the 16 pathogenic control variants with detectable current density (Z > −5), seven had abnormal Δ*V*_0.5_ act outside the blue normal range: V1392M, I1708T, T500M, E101K, R1348Q, Y1383C and D1643N. Hyperpolarising shifts in Δ*V*_0.5_ act with Z-scores <−3 were interpreted as GOF effects, whereas depolarising shifts with Z-scores > 3 were interpreted as LOF effects. Δ*V*_0.5_ act could not be measured for the remaining 14 pathogenic control variants because of severely reduced current amplitude. Data are presented as mean ± 95% CI.

The voltage-dependence of activation for the 26 benign and 16 pathogenic variants was represented as Δ*V*_0.5_ act, calculated as the variant *V*_0.5_ act minus the WT Δ*V*_0.5_ act recorded on the same plate (Fig. 4C). Based on the benign controls distribution, the mean Δ*V*_0.5_ act was 1.67 ± SD of 1.193 mV), and the normal functional range was defined as values within ±3 Z-score, shown by the blue shaded box in Fig. 4C. Of the 11 pathogenic variants with normal current density (Supplementary Table 2 and Fig. 3C), six showed a hyperpolarising shift in voltage-dependent of activation (Z < −3), consistent with GOF (E101K, T500M, R1348Q, Y1383C, V1392M and I1708T), and one showed a depolarising shift (Z > 3), consistent with LOF (D1643N). The remaining four variants (R279C, G297R, R1345Q and R1351L) had normal voltage-dependence of activation.

### Some pathogenic variants alter the voltage-dependence of inactivation

Cav2.1 inactivation was investigated by analysing peak currents recorded at zero mV after pre-conditioning pulses at voltages ranging from −90 to +40 mV (Fig. 5A). The midpoint of channel inactivation (*V*_0.5_ inact) was obtained by fitting a Boltzmann function to the envelope of peak currents (Fig. 5B). After subtracting the WT value recorded on the same plate, the mean Δ*V*_0.5_ inact for benign variants was 1.31 ± 3.221 mV, giving a ±3 Z-score normal range of −8.35 to +10.97 mV (Fig. 5C). No pathogenic variants showed a depolarising shift in steady-state inactivation, whereas four pathogenic variants showed a hyperpolarising shift in Δ*V*_0.5_ inact (V1392M, T500M, E101K and S218L), consistent with LOF (Fig. 5C). The four pathogenic variants that were not identified as abnormal by either current density or Δ*V*_0.5_ act (R1345Q, R279C, G297R and R1351L) had normal Δ*V*_0.5_ inact.

**Figure 5.**
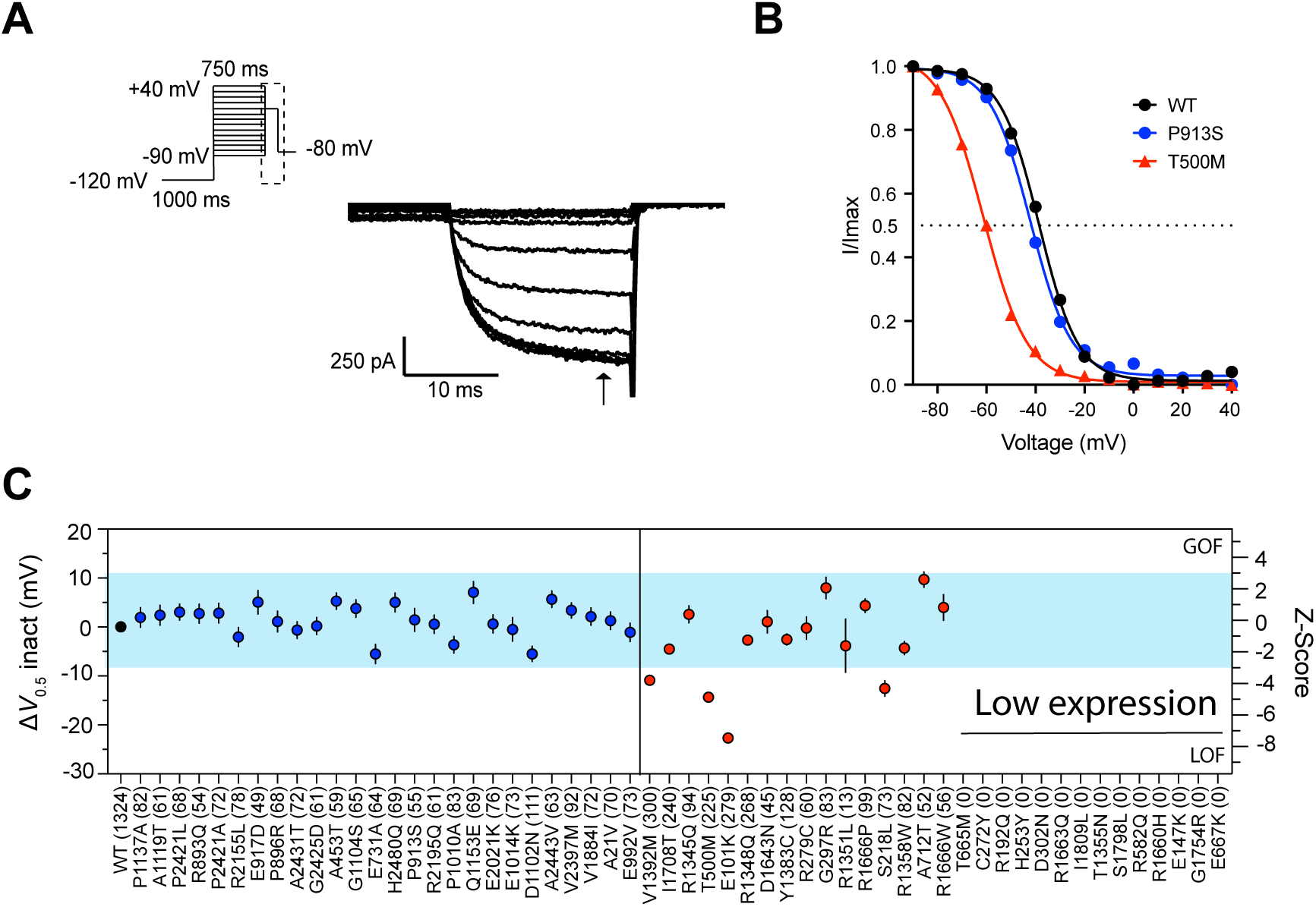
Benign control-calibrated assessment of voltage dependence of inactivation (*V*_0.5_ inact) identifies LOF *CACNA1A* variants. (A) The steady-state inactivation protocol consisted of 750 ms conditioning pulses from −90 mV to +40 mV in 10 mV increments, followed by a 20 ms test pulse to 0 mV from an inter-pulse holding potential of −120 mV to measure the fraction of available current. (B) Peak test-pulse currents were normalised to the maximum current and plotted against conditioning voltage. Solid lines represent Boltzmann fits to the experimental data. Example curves are shown for WT *CACNA1A* (*V*_0.5_ inact = −38.6 mV), the benign control P913S (*V*_0.5_ inact = −42 mV, blue), and the pathogenic control T500M (*V*_0.5_ inact = −61.2 mV, red), which shows a hyperpolarising shift consistent with loss of function. (C) The *V*_0.5_ inact of each variant was averaged and subtracted from the WT mean to calculate Δ*V*_0.5_ inact. Numbers of cells analysed for each variant are shown in parentheses. From analysis of WT cells, a minimum of 10 cells was required to detect a three Z-score difference in *V*_0.5_ inact at 90% power with a 95% CI. Using the Z-score grading system, Δ*V*_0.5_ inact values for all 26 benign *CACNA1A* control variants fell within the ±3 Z-score normal range (blue region). Four of 16 pathogenic control variants with detectable current density (Z > −5) had abnormal Δ*V*_0.5_ inact outside the blue normal range: V1392M, T500M, E101K and S218L. Hyperpolarising shifts in Δ*V*_0.5_ inact with Z-scores < −3 were interpreted as loss-of-function effects, whereas depolarising shifts with Z-scores > 3 were interpreted as gain-of-function effects. Δ*V*_0.5_ inact could not be measured for the remaining 14 pathogenic control variants because of severely reduced current amplitude. Data are presented as mean ± 95% CI.

### Pathogenic variants have mixed effects on recovery from inactivation

Recovery from inactivation (RFI) was investigated using a double-step protocol consisting of a pre-pulse to zero mV, followed by a step to –120 mV for intervals ranging from 50-ms to 15-s, and then a test pulse to zero mV (Fig. 6A). Current responses from test pulses, normalised to the maximum current obtained during the pre-pulse, were plotted versus the inter-pulse intervals and fitted with a double exponential function to obtain values for *T*_0.5_ RFI (Fig. 6B). In the example shown, WT has a *T*_0.5_ RFI of 2143 ms. For the benign controls, the normal range based on ±3 Z-score for *T*_0.5_ RFI normalised to WT, was 0.726 to 1.1 (Fig. 6C). Of the four pathogenic variants that failed to be identified as abnormal for current density, *V*_0.5_ act and *V*_0.5_ inact, three (R1345Q, G297R and R1351L) were found to have an increased *T*_0.5_ RFI, i.e. LOF that leaves R279C as the only pathogenic control variant that was functionally normal for all four parameters tested.

**Figure 6.**
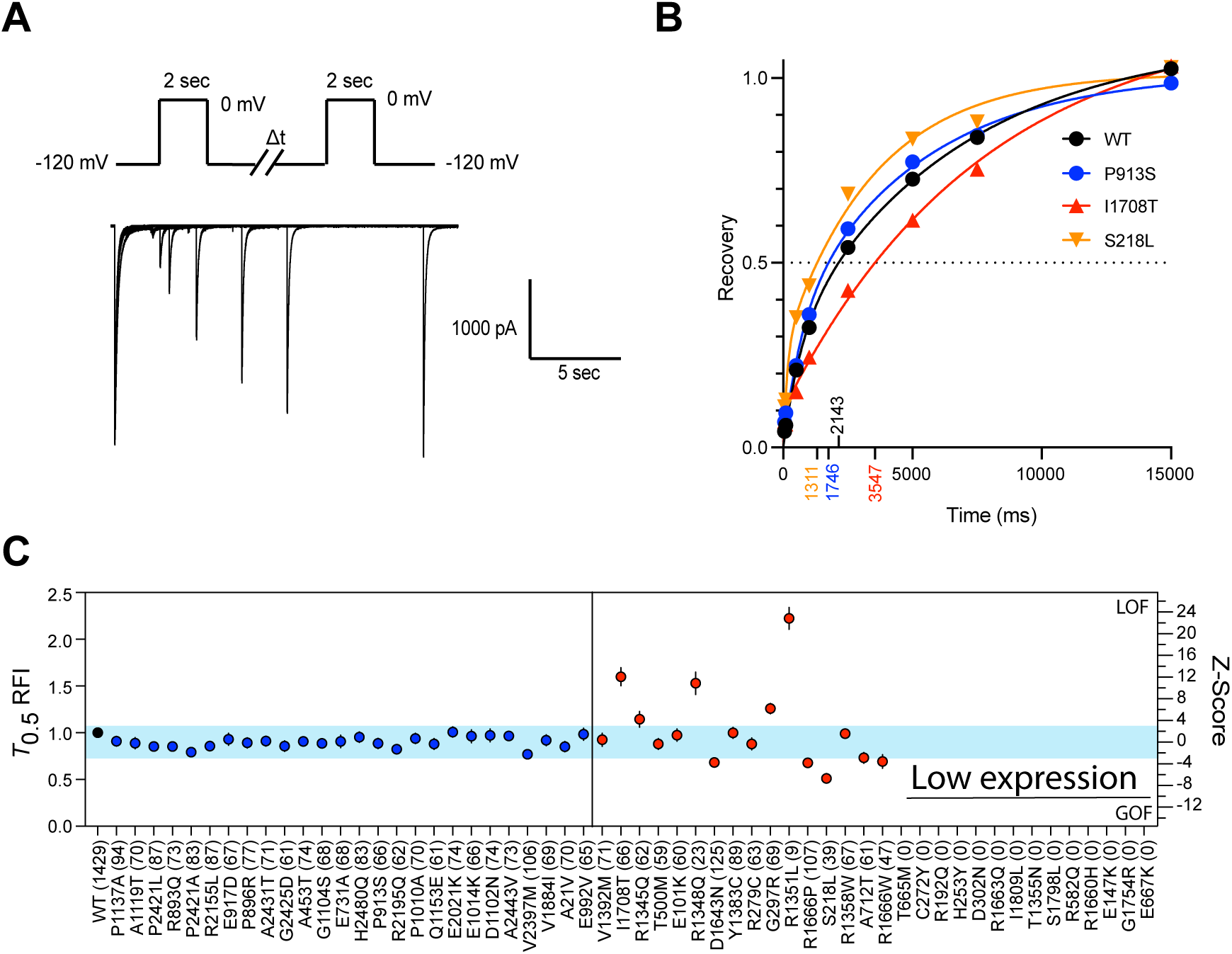
Establishment of a normal functional range for recovery from inactivation. (A) Recovery from inactivation (RFI) was assessed using a double-pulse protocol consisting of a 2-s pre-pulse to 0 mV, followed by recovery intervals at −120 mV ranging from 50 ms to 15 s, and then a 2-s test pulse to 0 mV. Representative traces are shown for a WT *CACNA1A* cell. Test-pulse currents were normalised to the maximum current during the corresponding pre-pulse. (B) The time required for 50% current recovery (*T*_0.5_ RFI) was determined by fitting the recovery time course with a double-exponential function. Example recovery curves are shown for WT *CACNA1A* (*T*_0.5_ RFI = 2143 ms), the benign control P913S (*T*_0.5_ RFI = 1746 ms), and pathogenic controls I1708T (*T*_0.5_ RFI = 3547 ms) and S218L (*T*_0.5_ RFI = 1311 ms). (C) Numbers of cells analysed for each variant are shown in parentheses. From analysis of 1040 WT cells, a minimum of 37 cells was required to detect a three Z-score difference in *T*_0.5_ RFI at 90% power with a 95% CI. Using the Z-score grading system, the RFI values for all 26 benign *CACNA1A* controls fell within the ±3 Z-score normal range (blue region). Eight of 16 pathogenic control variants with detectable current density (Z > −5 in Fig. 3) had abnormal *T*_0.5_ RFI: I1708T, R1345Q, R1348Q, G297R, R1351L, R1666P, S218L and R1666W. Among these, three variants had normal current density, *V*_0.5_ act and *V*_0.5_ inact. Z-scores < −3 indicate faster recovery from inactivation, consistent with a gain-of-function effect, whereas Z-scores > 3 indicate slower recovery from inactivation, consistent with a loss-of-function effect. *T*_0.5_ RFI could not be measured for 14 pathogenic control variants with current density Z-scores < −5 because of severely reduced current amplitude. Data are presented as mean ± 95% CI.

### Calibrated Z-score thresholds provide strong functional evidence for classification

Before applying functional data from this patch-clamp assay to clinical *CACNA1A* variant classification, the assay’s performance was assessed in accordance with the recommendations of the ClinGen Sequence Variant Interpretation Working Group.^22^ The range for normal Cav2.1 function was defined as ±3 Z-score of the mean of the 26 benign variants for each of the four parameters tested: current density, Δ*V*_0.5_ act, Δ*V*_0.5_ inact and *T*_0.5_ RFI. Among the 30 pathogenic controls, 29 had at least one abnormal parameter (Supplementary Fig. 1A). This yielded likelihood ratios of 0.033 for normal function and 25.13 for abnormal function (Supplementary Fig. 1B), corresponding to strong benign functional evidence (BS3) and strong pathogenic functional evidence (PS3), respectively.^22^

### *CACNA1A* variants from Australian patients

A total of 23 *CACNA1A* variants from Australian patients were functionally assessed using the calibrated *CACNA1A* assay (Fig. 7A). Of these 23 variants, 19 were classified as VUS, two as likely pathogenic, and two as pathogenic before application of functional evidence (Fig. 7F). Among the 11 VUS with abnormal function in at least one parameter, nine showed LOF in current density: R588C, I613M, Y692C, P1218S, K1437Del, D1443G, P1692L, G1844D, V2440I (Fig. 7B); two cause LOF by altering *V*_0.5_ act: I141T and R588C (Fig. 7C); three show a GOF effect in *T*_0.5_ RFI: I141T, R369Q and G1844D (Fig. 7D), and one has an GOF effect in *V*_0.5_ inact: R588C (Fig. 7E). Of these 11 functionally abnormal VUS, ten were reclassified as LP following application of abnormal functional evidence, while V2440I remained classified as VUS (Fig. 7F, Supplementary Table 1). The clinical phenotypes of the abnormal *CACNA1A* variants were heterogeneous and predominantly neurodevelopmental, including global developmental delay, intellectual disability, delayed speech and language development, hypotonia, autism spectrum disorder, and congenital or dysmorphic features in several individuals (Supplementary Table 3). Epilepsy-related presentations included absence seizures, myoclonic seizures, focal motor or generalised tonic-clonic seizures, status epilepticus, infantile spasms and epileptic encephalopathy, with drug-resistant epilepsy documented in selected cases. Cerebellar and motor features were also common, including episodic or progressive ataxia, nystagmus, tremor, impaired walking and cerebellar atrophy, while neuroimaging abnormalities included ventriculomegaly, cortical dysplasia, lissencephaly, corpus callosum abnormality and white matter changes suspicious for leukodystrophy.

**Figure 7.**
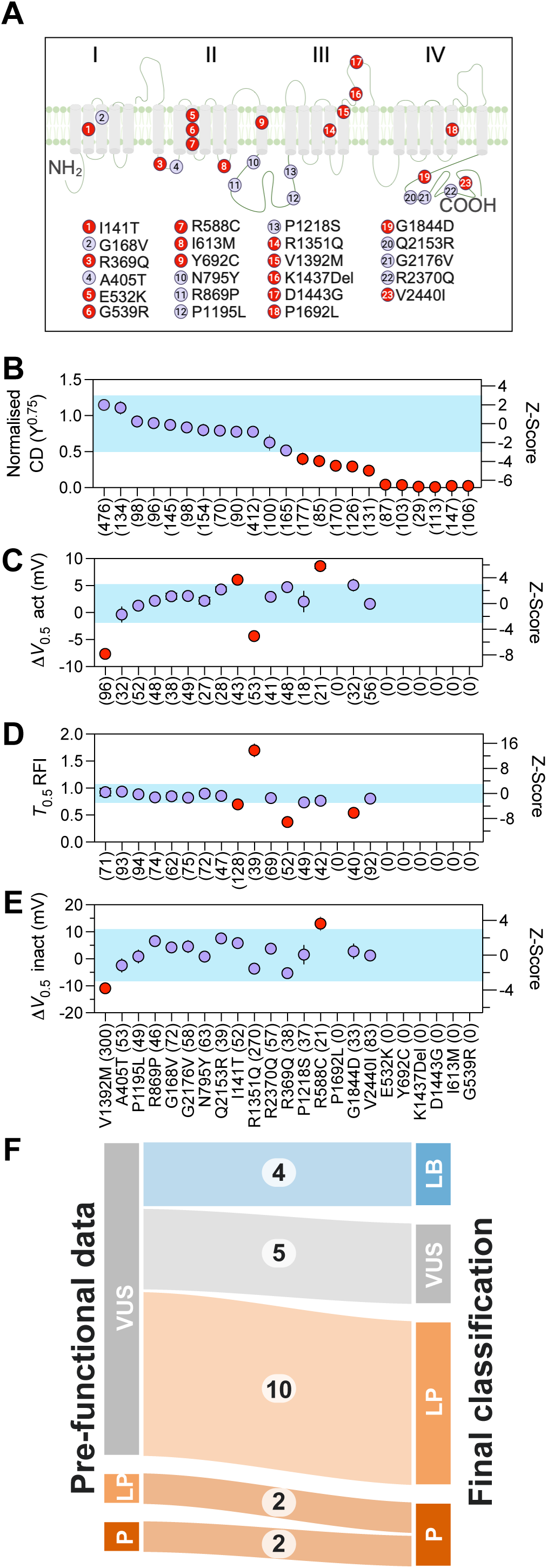
Functional characterisation and classification of *CACNA1A* variants identified in Australian patients. (A) Topological positions of the Australian patient *CACNA1A* variants analysed in this study. Variants with abnormal function in at least one electrophysiological parameter are indicated as red. (B) Normalised current density. (C) Δ*V*_0.5_ act. (D) *T*_0.5_ RFI. (E) Δ*V*_0.5_ inact. For panels B–E, values within the ±3 Z-score range were considered functionally normal, whereas values outside this range were considered abnormal. Data are presented as mean ± 95% CI. (F) Sankey plot showing variant classification before and after application of calibrated functional evidence. Of 19 variants initially classified as VUS, ten were reclassified as likely pathogenic, four were reclassified as likely benign and five remained VUS. Two variants initially classified as likely pathogenic were upgraded to pathogenic, while two variants initially classified as pathogenic remained pathogenic. See Supplementary Table 1 for classification details.

## Discussion

We developed a high-throughput automated patch-clamp assay for *CACNA1A* variants that meets ClinGen recommendations for functional evidence by incorporating a calibrated control framework. Using 26 benign variants, we defined benign control-calibrated normal ranges for four electrophysiological parameters: current density, Δ*V*_0.5_ act, Δ*V*_0.5_ inact and *T*_0.5_ RFI. With Z = ±3 as the threshold for normal function, all benign controls remained within the normal range, whereas 29 of 30 pathogenic controls were abnormal in at least one parameter, yielding 100% specificity and 96.7% sensitivity. Current density was the primary discriminator, identifying 19 pathogenic controls with LOF, while gating and recovery from inactivation analyses identified additional variants with abnormal function. Applying this calibrated framework to 19 VUS identified in Australian patients, we found 11 with functional abnormalities, of which ten were reclassified as LP after integrating PS3 evidence with other ACMG/AMP criteria.

### VUS burden in *CACNA1A*

The topological distribution of ClinVar missense variants further highlights the need for functional evidence. Although P/LP variants were relatively enriched in transmembrane regions, VUS represented the majority of missense variants across intracellular, transmembrane and extracellular regions (Supplementary Fig. 2). This indicates that variant location alone is insufficient for classification, particularly because many VUS occur outside regions that are obviously enriched for P/LP variation. Therefore, a calibrated functional assay is needed to distinguish variants with normal Cav2.1 function from those with abnormal current density, gating or recovery from inactivation.

### Clinical interpretation of functional evidence

According to the ClinGen recommendations, benign and pathogenic variant controls are essential for establishing the specificity and sensitivity of any functional assay. For this *CACNA1A* patch-clamp assay, 26 benign controls were used to define the biological normal range and its variance. This approach facilitates objective differentiation between variants with clearly abnormal function and variants that fall within the benign-calibrated functional range. Simultaneously, the inclusion of pathogenic controls enables assay sensitivity to be estimated and supports application of abnormal results as evidence for variant classification. By utilising the benign distribution to develop Z-score metrics for current density, Δ*V*_0.5_ act, Δ*V*_0.5_ inact and *T*_0.5_ RFI, the method enables parameter-specific assessments of functional normality or abnormality, which is important because *CACNA1A* variants can cause LOF, GOF or mixed phenotypes. This, in turn, elucidates variant-specific defects for each VUS, acknowledging that not all variants will exhibit abnormalities across every metric and that severe LOF in current density will preclude measurement of gating parameters. The Z-score metric has previously been utilised in inherited cardiac channelopathies to provide clinically interpretable evidence suitable for routine evaluation of VUS.^29,30^ In this *CACNA1A* patch-clamp assay, a Z-score threshold of ±3 has been used as the criterion for strong functional evidence, which minimises false-positive results but may reduce assay sensitivity for mild LOF/GOF variants. By using four biophysical parameters, the assay achieved 100% specificity and 96.7% sensitivity.

### Functionally abnormal VUS identified in Australian patients

Subsequently, we employed this functional assay to evaluate *CACNA1A* variants identified in Australian patients. Of the 23 variants assessed, four were already classified as pathogenic or likely pathogenic before application of functional evidence, and all four were detected as abnormal by at least one electrophysiological parameter. This is noteworthy because functional evidence represents only one component of the ACMG/AMP framework, and variant classification relies on multiple independent lines of evidence. Of the 19 VUS, the assay identified 11 as functionally abnormal. Nine VUS showed LOF in current density: R588C, I613M, Y692C, P1218S, K1437Del, D1443G, P1692L, G1844D and V2440I. Two VUS showed abnormal Δ*V*_0.5_ activation: I141T and R588C. Three VUS showed GOF effects in *T*_0.5_ RFI: I141T, R369Q and G1844D, and one VUS, R588C, showed a GOF effect in Δ*V*_0.5_ inactivation. Following application of abnormal functional evidence as PS3, ten of the 11 functionally abnormal VUS were reclassified as likely pathogenic, whereas V2440I remained classified as VUS.

### Biophysical heterogeneity of *CACNA1A* variant function

Our findings are broadly consistent with the recent large-scale functional study of *CACNA1A* missense variants by Kurganov and colleagues,^31^ despite differences in assay platform, variant selection and study aims. Seven benign variants were shared between studies (A453T, E731A, E917D, E992V, P1010A, E1014K and G1104S), and all were functionally normal in both datasets. There was also broad agreement in the direction of effect for several shared pathogenic variants, including GOF activation shifts for E101K, R1348Q, Y1383C, V1392M and I1708T, and reduced current density for R1358W. The key distinction of the present study is the use of a benign and pathogenic control-calibrated framework to define assay-specific normal ranges, estimate likelihood ratios, and convert Cav2.1 biophysical measurements into ACMG/AMP-compatible functional evidence.

However, this concordance should be interpreted at the mechanistic rather than the individual variant level, as *CACNA1A* is pleiotropic and individual variants do not always map cleanly to a single EA2, FHM1 or neurodevelopmental presentation. In our assay, reduced current density was observed in more than half of the pathogenic variants. Pathogenic variants with partial current loss frequently showed GOF changes in activation and/or recovery from inactivation, and some pathogenic variants with normal current density exhibited abnormal gating. These observations reinforce the view that biophysical direction alone does not rigidly determine phenotype, and that partial reduction in macroscopic current does not exclude gating changes that may increase channel availability under specific stimulation conditions. Such mixed effects may help explain why some *CACNA1A* variants do not segregate cleanly into EA2 or FHM1, but instead produce overlapping clinical features, including ataxia with migraine.

Although the heterologous HEK293 patch-clamp assay is highly scalable for studying large numbers of variants, it has inherent limitations for interpreting *CACNA1A* biology in its full physiological context. HEK293 cells do not recapitulate the complex presynaptic architecture, neuromodulatory environment, auxiliary subunit diversity, splice-isoform context, or developmental state of native neurons, and therefore cannot fully model how altered Cav2.1 biophysics affects synaptic release and network excitability. There was one pathogenic control, R279C, that remained within the benign-calibrated normal range across all four measured parameters, indicating that normal function in this assay does not exclude all possible pathogenic mechanisms. More physiologically relevant systems, such as iPSC-derived neuronal models, may provide complementary insights into selected variants, although these approaches have their own limitations related to maturation, variability, scalability, and control-based calibration. Because the primary aim of this study was to establish a robust assay for variant interpretation rather than to model downstream neuronal consequences, a calibrated HEK293-based assay using extensive benign and pathogenic controls remains aligned with ClinGen recommendations for generating clinically interpretable functional evidence. Within this framework, abnormal function can support PS3 and normal function can support BS3; however, normal functional evidence BS3 was applied conservatively at moderate strength when integrated with other ACMG/AMP criteria for classifying *CACNA1A* variants.

## Conclusion

*CACNA1A* missense variants can produce LOF, GOF or mixed functional effects. We developed a benign control-calibrated automated patch-clamp assay that integrates current density, Δ*V*_0.5_ activation, Δ*V*_0.5_ inactivation and *T*_0.5_ recovery from inactivation to distinguish functionally normal from abnormal variants. This calibrated framework converts Cav2.1 biophysical measurements into clinically interpretable PS3 for abnormal function and BS3 for normal function, thereby supporting VUS reclassification in *CACNA1A*.

## Data Availability

All data produced in the present study are available upon reasonable request to the authors

## Acknowledgement

We acknowledge the support from the Victor Chang Cardiac Research Institute Innovation Center, funded by the New South Wales Government, Australia. This research was funded by a Medical Research Future Fund: Genomics Health Futures Mission grant MRF2016760 (CAN/JIV) and NHMRC Investigator Grant App2041619 (JIV).

## Ethics

The study was conducted in accordance with the ethical principles of the Declaration of Helsinki (2013 revision). Ethics approval was granted by the Human Research Ethics Committee of the Royal Children’s Hospital, Melbourne (approval number 125558). Analyses were performed using publicly available genetic datasets generated during routine clinical genetic testing, for which appropriate patient consent had been obtained by the originating clinical providers.

