## Supplemental Table 2 for "A benign control-calibrated multiparametric functional assay supports *CACNA1A* variant assessment"

**Table S2.** **Z-scores for multi-parameter electrophysiological characterisation.**

|  | Variant | Z-score Current density | Z-score *V*_0.5_ act | Z-score *V*_0.5_ inact | Z-score *T*_0.5_ RFI | Location |
| --- | --- | --- | --- | --- | --- | --- |
| WT | CACNA1A_WT | 0.86 | -1.38 | -0.41 | 1.76 | #N/A |
| Benign | P1137A | 1.90 | -1.14 | 0.19 | 0.18 | Cytoplasmic |
| Benign | A1119T | 1.68 | -0.80 | -0.35 | -0.17 | Cytoplasmic |
| Benign | P2421L | 1.58 | 0.35 | 0.53 | -0.80 | Cytoplasmic |
| Benign | R893Q | 1.39 | 0.51 | 0.43 | -0.78 | Cytoplasmic |
| Benign | P2421A | 0.95 | 1.90 | 0.46 | -1.84 | Cytoplasmic |
| Benign | R2155L | 0.66 | -0.77 | -1.05 | -0.70 | Cytoplasmic |
| Benign | E917D | 0.57 | 0.58 | 1.16 | 0.55 | Cytoplasmic |
| Benign | P896R | 0.43 | -0.36 | -0.07 | -0.11 | Cytoplasmic |
| Benign | A2431T | 0.39 | 0.16 | -0.61 | 0.22 | Cytoplasmic |
| Benign | G2425D | 0.38 | 0.10 | -0.35 | -0.70 | Cytoplasmic |
| Benign | A453T | 0.28 | 0.70 | 1.23 | 0.14 | Cytoplasmic |
| Benign | G1104S | 0.07 | 0.09 | 0.77 | -0.20 | Cytoplasmic |
| Benign | E731A | -0.02 | -1.55 | -2.13 | 0.16 | Cytoplasmic |
| Benign | H2480Q | -0.05 | 1.27 | 1.15 | 0.95 | Cytoplasmic |
| Benign | P913S | -0.06 | -0.03 | 0.04 | -0.22 | Cytoplasmic |
| Benign | R2195Q | -0.19 | 1.18 | -0.23 | -1.30 | Cytoplasmic |
| Benign | P1010A | -0.35 | -1.92 | -1.54 | 0.71 | Cytoplasmic |
| Benign | Q1153E | -0.55 | 1.33 | 1.78 | -0.31 | Cytoplasmic |
| Benign | E2021K | -0.64 | -0.71 | -0.22 | 1.89 | Cytoplasmic |
| Benign | E1014K | -0.74 | -0.34 | -0.57 | 1.11 | Cytoplasmic |
| Benign | D1102N | -0.96 | -1.43 | -2.12 | 1.26 | Cytoplasmic |
| Benign | A2443V | -0.97 | 1.06 | 1.35 | 1.15 | Cytoplasmic |
| Benign | V2397M | -1.06 | -0.27 | 0.64 | -2.22 | Cytoplasmic |
| Benign | V1884I | -1.35 | -0.35 | 0.25 | 0.38 | Cytoplasmic |
| Benign | A21V | -1.59 | -0.90 | -0.01 | -0.81 | Cytoplasmic |
| Benign | E992V | -1.77 | 1.34 | -0.75 | 1.48 | Cytoplasmic |
| Pathogenic | V1392M | 1.99 | -7.81 | -3.79 | 0.46 | S5 of repeat III |
| Pathogenic | I1708T | 0.66 | -6.18 | -1.81 | 12.15 | S5 of repeat IV |
| Pathogenic | R1345Q | -0.29 | -2.12 | 0.39 | 4.26 | S4 of repeat III |
| Pathogenic | T500M | -0.67 | -4.60 | -4.86 | -0.31 | S1 of repeat II |
| Pathogenic | E101K | -0.78 | -9.98 | -7.45 | 1.32 | S1 of repeat I |
| Pathogenic | R1348Q | -1.20 | -4.69 | -1.24 | 10.92 | S4 of repeat III |
| Pathogenic | D1643N | -2.49 | 7.10 | -0.08 | -3.80 | S3 of repeat IV |
| Pathogenic | Y1383C | -2.51 | -8.30 | -1.20 | 1.75 | S5 of repeat III |
| Pathogenic | R279C | -2.57 | 0.11 | -0.47 | -0.30 | Extracellular |
| Pathogenic | G297R | -2.67 | 2.24 | 2.07 | 6.23 | Extracellular |
| Pathogenic | R1351L | -2.94 | -0.42 | -1.60 | 22.96 | S4 of repeat III |
| Pathogenic | R1666P | -3.48 | -4.75 | 0.94 | -3.80 | S4 of repeat IV |
| Pathogenic | S218L | -3.57 | -4.64 | -4.31 | -6.69 | Cytoplasmic |
| Pathogenic | R1358W | -3.64 | -6.22 | -1.75 | 1.61 | Cytoplasmic |
| Pathogenic | A712T | -3.74 | -3.58 | 2.59 | -2.89 | S6 of repeat II |
| Pathogenic | R1666W | -4.54 | 0.38 | 0.83 | -3.57 | S4 of repeat IV |
| Pathogenic | T665M | -5.52 | N/A | N/A | N/A | Extracellular |
| Pathogenic | C272Y | -5.53 | N/A | N/A | N/A | Extracellular |
| Pathogenic | R192Q | -5.58 | N/A | N/A | N/A | S4 of repeat I |
| Pathogenic | H253Y | -5.80 | N/A | N/A | N/A | Extracellular |
| Pathogenic | D302N | -5.88 | N/A | N/A | N/A | Extracellular |
| Pathogenic | R1663Q | -5.92 | N/A | N/A | N/A | S4 of repeat IV |
| Pathogenic | I1809L | -6.32 | N/A | N/A | N/A | Cytoplasmic |
| Pathogenic | T1355N | -6.39 | N/A | N/A | N/A | S4 of repeat III |
| Pathogenic | S1798L | -6.51 | N/A | N/A | N/A | S6 of repeat IV |
| Pathogenic | R582Q | -6.57 | N/A | N/A | N/A | S4 of repeat II |
| Pathogenic | R1660H | -6.60 | N/A | N/A | N/A | S4 of repeat IV |
| Pathogenic | E147K | -6.61 | N/A | N/A | N/A | S2 of repeat I |
| Pathogenic | G1754R | -6.61 | N/A | N/A | N/A | Extracellular |
| Pathogenic | E667K | -6.63 | N/A | N/A | N/A | Extracellular |
| Patient | V1392M | 1.99 | -7.81 | -3.79 | 0.46 | S5 of repeat III |
| Patient | A405T | 1.69 | -1.68 | -1.17 | 0.61 | Cytoplasmic |
| Patient | P1195L | 0.24 | -0.33 | -0.14 | -0.19 | Cytoplasmic |
| Patient | R869P | 0.06 | 0.45 | 1.63 | -1.25 | Cytoplasmic |
| Patient | G168V | -0.12 | 1.14 | 0.91 | -0.86 | S3 of repeat I |
| Patient | G2176V | -0.39 | 1.20 | 1.01 | -1.27 | Cytoplasmic |
| Patient | N795Y | -0.68 | 0.44 | -0.14 | 0.00 | Cytoplasmic |
| Patient | Q2153R | -0.73 | 2.20 | 1.93 | -0.75 | Cytoplasmic |
| Patient | I141T | -0.84 | 3.69 | 1.41 | -3.45 | S2 of repeat I |
| Patient | R1351Q | -0.85 | -5.42 | -1.53 | 13.83 | S4 of repeat III |
| Patient | R2370Q | -2.00 | 1.05 | 0.76 | -1.41 | Cytoplasmic |
| Patient | R369Q | -2.84 | 2.54 | -2.06 | -9.00 | Cytoplasmic |
| Patient | P1218S | -3.74 | 0.54 | 0.07 | -2.86 | Cytoplasmic |
| Patient | R588C | -3.96 | 5.81 | 3.63 | -2.30 | S4 of repeat II |
| Patient | P1692L | -4.46 | N/A | N/A | N/A | S5 of repeat IV |
| Patient | G1844D | -4.54 | 2.52 | 0.46 | -6.20 | Cytoplasmic |
| Patient | V2440I | -4.97 | -0.05 | -0.02 | -1.54 | Cytoplasmic |
| Patient | E532K | -6.47 | N/A | N/A | N/A | S2 of repeat II |
| Patient | Y692C | -6.50 | N/A | N/A | N/A | S6 of repeat II |
| Patient | I613M | -6.59 | N/A | N/A | N/A | Cytoplasmic |
| Patient | G539R | -6.61 | N/A | N/A | N/A | S2 of repeat II |
| Patient | K1437del | -6.68 | N/A | N/A | N/A | Extracellular |
| Patient | D1443G | -6.71 | N/A | N/A | N/A | Extracellular |
| Negative | Negative | -6.70 | N/A | N/A | N/A | #N/A |

Electrophysiological parameters with normal function are indicated by a Z-score within ± 3 (blue), whereas abnormal function is indicated with a Z-score outside the ±3 (red: LOF, peach: GOF). A variant is considered as functionally normal only if all its measured electrophysiological parameters are within ±3 Z-score (blue). V1392M is also one of the pathogenic controls.
