## Supplemental Figure 1 for "A benign control-calibrated multiparametric functional assay supports *CACNA1A* variant assessment"

**A**

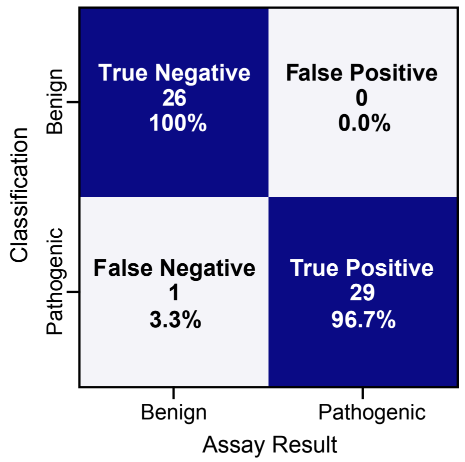


**B**

| *CACNA1A* assay | Likelihood Ratio | ACMG Evidence strength |
|---|---|---|
| Normal Function | 0.033 | BS3 |
| Abnormal function | 25.13 | PS3 |
